# Harnessing Deep Learning to Detect Bronchiolitis Obliterans Syndrome from Chest CT

**DOI:** 10.1101/2024.02.07.24302414

**Authors:** Mateusz Kozinski, Doruk Oner, Jakub Gwizdala, Catherine Beigelman, Pascal Fua, Angela Koutsokera, Alessio Casutt, Michele De Palma, John-David Aubert, Horst Bischof, Christophe von Garnier, Sahand Rahi, Martin Urschler, Nahal Mansouri

**Affiliations:** Institute of computer graphics and vision, Technische Universität Graz, Austria; Computer Vision Laboratory, School of Computer and Communication Sciences – IC, École Polytechnique Fédérale de Lausanne (EPFL), Lausanne, Switzerland; Department of Diagnostic and Interventional Radiology, Lausanne University Hospital (CHUV), Lausanne, Switzerland; Division of Pulmonary Medicine, Department of Medicine, Lausanne University Hospital (CHUV), University of Lausanne (UNIL), Lausanne, Switzerland; Transplantation Center, Lausanne University Hospital (CHUV) and University of Lausanne (UNIL), Lausanne, Switzerland; Swiss Institute for Experimental Cancer Research (ISREC), School of Life Sciences, École Polytechnique Fédérale de Lausanne (EPFL), Lausanne, Switzerland; Laboratory of the Physics of Biological Systems, Institute of Physics, École Polytechnique Fédérale de Lausanne (EPFL), Lausanne, Switzerland; Institute for Medical Informatics, Statistics and Documentation, Medical University of Graz, Austria

## Abstract

Bronchiolitis Obliterans Syndrome (BOS), a fibrotic airway disease following lung transplantation, conventionally relies on pulmonary function tests (PFTs) for diagnosis due to limitations of CT images. Thus far, deep neural networks (DNNs) have not been used for BOS detection. We optimized a DNN for detection of BOS solely using CT scans by integrating an innovative co-training method for enhanced performance in low-data scenarios. The novel auxiliary task is to predict the temporal precedence of CT scans of BOS patients. We tested our method using CT scans at various stages of inspiration from 75 post-transplant patients, including 26 with BOS. The method achieved a ROC-AUC of 0.90 (95% CI: 0.840-0.953) in distinguishing BOS from non-BOS CT scans. Performance correlated with disease progression, reaching 0.88 ROC-AUC for stage I, 0.91 for stage II, and an outstanding 0.94 for stage III BOS. Importantly, performance parity existed between standard and high-resolution scans. Particularly noteworthy is the DNN’s ability to predict BOS in at-risk patients (FEV1 between 80% and 90% of best FEV1) with a robust 0.87 ROC-AUC (CI: 0.735-0.974). Using techniques for visually interpreting the results of deep neural networks, we reveal that our method is especially sensitive to hyperlucent areas compatible with air-trapping or bronchiectasis. Our approach shows the potential to improve BOS diagnosis, enabling early detection and management. Detecting BOS from low-resolution scans reduces radiation exposure and using scans at any stage of respiration makes our method more accessible. Additionally, we demonstrate that techniques that limit overfitting are essential to unlocking the power of DNNs in scenarios with scarce training data. Our method may enable clinicians to use DNNs in studies where only a modest number of patients is available.

## 1. Introduction

Bronchiolitis obliterans syndrome (BOS) is a progressive fibrotic lung disease affecting lung transplantation recipients and patients with hematopoietic transplantation ^1,2^. In lung transplant recipients, BOS is defined as graft deterioration secondary to progressive airway disease for which there is no other cause. The severity of BOS is defined by the decline of forced expiratory volume in the first second (FEV_1_) from the patient’s best value ^3^. BOS is associated with poor overall survival (median of 2.5 years after lung transplantation). Unfortunately, the alterations in spirometry are only apparent after the establishment of airway fibrosis. At this point, the disease is irreversible with no curative treatment ^4^. Consequently, early diagnosis is pivotal for optimal patient management and follow-up ^5^.

The diagnosis of BOS presents multiple challenges. Surgical lung biopsies are not routinely performed due to the high risk of complications in lung transplant recipients ^6,7^. Less invasive, transbronchial biopsies have a low diagnostic sensitivity. The clinical presentation of BOS is similar to other obstructive lung diseases including asthma, COPD, and bronchiectasis, which may also present with an irreversible obstructive pattern on spirometry ^6^. Consequently, current diagnostic methods centered on pulmonary function tests (PFTs), e.g. decline in FEV_1_, lack specificity, and chest imaging for diagnosing BOS is not fully established. Typical findings, such as parietal wall bronchial thickening and mosaic attenuation/perfusion pattern with air-trapping on expiration, can only be seen with advanced disease, and some BOS cases remain difficult to diagnose even in advanced forms. Moreover, CT scans are only considered helpful if performed at high resolution and preferably during the expiratory phase of breathing, which conveys higher radiation exposure and maneuvers that may not be feasible for patients with poor lung capacity ^8,9^.

Ongoing research on computational methods to detect BOS from CT scans were designed to address the current limitations of CT imaging in BOS diagnosis. However, previous methods ^10,11,12,13,14,15,16,17,18^ rely on heavily engineered procedures for extracting information from the scans, and designing these procedures to maximize robustness in scans taken at different resolutions or respiration phases is not feasible. These drawbacks can be overcome by using a deep neural network (DNN) ^19^, which have thus far not been used for BOS detection. By contrast to previous visual recognition approaches, DNNs learn to produce disease likelihoods directly from images. This end-to-end learning, from raw data to disease likelihood, allows DNNs to capture subtle manifestations of diseases in CT scans, including ones that might be too difficult to quantify with manually designed procedures, and gives them the flexibility needed to adapt to variation in scan appearance. However, DNNs require large volumes of training data to reach their full performance, which so far hindered their application to studies with only a small number of patients.

In this study, we developed an innovative DNN technique for BOS detection. Integrating a novel as well as multiple existing methods to enhance training data efficiency, we crafted a state-of-the-art approach that simultaneously trains the DNN to detect BOS and differentiate between late and early stages of the disease. Our DNN-based BOS detector is the first to detect BOS from standard-resolution scans taken at any stage of respiration. Moreover, it shows promise in predicting future BOS onset in patients at risk. Finally, our contribution goes beyond detecting BOS: Thanks to its data-efficiency, our method enables using DNNs in studies where the number of available patients is limited due to the low incidence of the targeted disease or by other constraints.

## Methods

### 1.1. Study participants

Approval for this study was obtained from the relevant ethics committee (CER-VD reference number 2020-02455). We initially included 130 lung transplant recipients presenting to University Hospital of Lausanne, Switzerland between 1990 and 2020. Among those, 45 patients were diagnosed with BOS. The 85 patients without BOS had either normal PFTs, restrictive lung disease, or obstructive lung diseases other than BOS, e.g., airway stenosis. For each patient, the presence or absence of BOS in chest imaging was confirmed by an expert thoracic radiologist and an expert pulmonologist. Patients with insufficient PFT documentation or missing CT scans were excluded from the analysis. We retained 75 patients, including 26 with confirmed BOS diagnosis (**Figure 1**).

**Figure 1:**
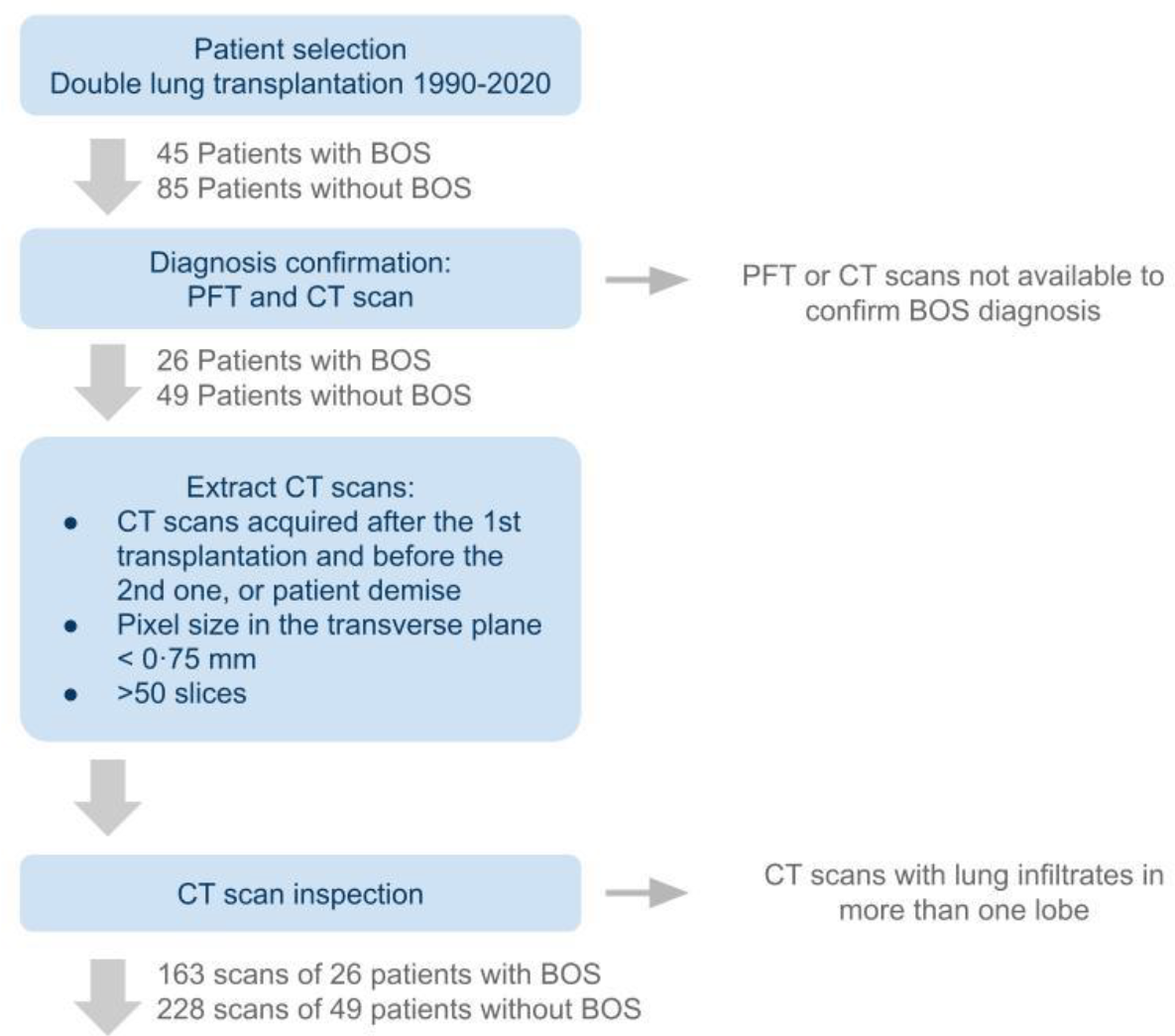
CT scan selection protocol.

For each of the selected patients, we extracted and anonymized thoracic CT scans from the hospital’s picture archiving and communication system (PACS). We incorporated CT scans from the time of transplantation to either the time of data extraction, the date of the patients’ demise or their second transplantation. Within the inclusion period, patients presented for a CT scan between one and eighteen times and the majority of patients had between three and ten visits (80%). Whenever PACS contained more than one copy of a scan, e.g., reconstructed with the standard kernel and the lung kernel, we retained all the copies. Scans with pixel size larger than 0·75 mm in the transverse plane and ones containing less than fifty transverse slices were excluded. Finally, we inspected the entire set of scans and removed those with lung infiltrates in more than one lobe. For patients not diagnosed with BOS, this approach resulted in 228 scans. Patients eventually diagnosed with BOS contributed a total of 163 scans, with 105 scans acquired after the onset of the disease. The onset was defined as the time when the FEV1 consistently fell below 80% of the patient’s best value, computed as the average of the two best measurements taken at least three weeks apart, in alignment with clinical guidelines ^10^.

### 1.2. Evaluation methodology

Our main objective was to evaluate the performance of the DNN in differentiating patients with BOS from patients without BOS. To that end, we analyzed the CT scans of BOS patients taken after their FEV_1_ fell below 80% of the best FEV_1_, which corresponds to the clinical criterion of BOS stage I ^20^, and all scans in patients never diagnosed with BOS (non-BOS).

To understand how the BOS detection accuracy changes as the disease progresses, we conducted evaluation on CT scans of BOS patients in stages I, II, and III of the disease, defined in accordance with the current diagnostic guidelines as a fall of FEV_1_ below 80%, 65%, and 50% of the best FEV_1_ on at least two consecutive tests taken no less than three weeks apart. ^20^ We evaluated the network’s ability to distinguish between these CT scans and CT scans of non-BOS patients. To enhance the versatility of our approach, we opted for CT scans acquired at any stage of inspiration, diverging from the conventional practice of relying on expiratory CT scans for BOS diagnosis.

From the clinical perspective, the added value of a method to automatically detect BOS from CT scans largely depends on its capacity to detect patients at risk of developing the disease before the lung function declines. To verify this capacity, we designed a comparison that closely mimics a realistic clinical scenario, in which a patient whose FEV_1_ has decreased but remains above the threshold of BOS stage I needs to be screened for the risk of developing BOS in the future. We evaluated the performance of the network in distinguishing the scans of BOS patients taken before the diagnosis with FEV_1_ between 80% and 90% of best FEV_1_ from the scans of non-BOS patients with relative FEV_1_ in the same range.

Since high-resolution scans (slice thickness below 1.25 mm) contain more information than standard-resolution ones, we also investigated if this difference affects the ability of the DNN to learn to extract diagnostic information. We verified this by re-training the DNN on data sets restricted to high-resolution scans and limiting the test sets accordingly.

### 1.3. Performance evaluation criteria and statistical analysis

We used the Chi-squared test for contingency tables to evaluate the match in sex between the BOS and non-BOS groups, and the Student’s t-test for independent samples to compare patient ages and FEV_1,_ measured 3 and 30 months after transplantation.

We performed five-fold cross-validation. We randomly divided the cohort of 26 patients with BOS and 49 non-BOS patients into four splits of 5 patients with BOS and 10 patients without BOS and one split of 6 patients with BOS and 9 patients without BOS. We trained our DNN five times according to the procedure described below, each time leaving scans of one split of patients out of the training set. We tested the resulting classifier on the scans that were not used for training.

We present the results using the receiver operating characteristic area under curve (ROC-AUC). When computing the curves, we weighted the results for individual scans to prevent biasing the results to patients who had scans performed more often. The details of this procedure are provided in the supplementary material.

To summarize the performance, we computed a single ROC curve for test predictions computed for CT scans of patients from all five splits and evaluated the corresponding AUC. We call this metric “aggregated AUC”. Unlike the average of AUC computed for individual splits, the aggregated AUC provides a conservative performance estimate, because each point in the aggregated ROC curve results from applying the same threshold to predictions produced by five independently trained DNNs with different optimal thresholds.

We could not apply the standard approach to computing confidence intervals for the aggregated ROC-AUC values since predictions produced by the DNN for two scans of the same patient are not statistically independent. We therefore resorted to bootstrapping by hierarchical case resampling ^21^. We used ten thousand simulations to estimate the 95% confidence intervals. See supplementary material section C1 for details. To additionally illustrate the variability of the performance of our method, we report the performance attained on individual splits in the form of box plots containing five ROC-AUC values, one for each training run.

### 1.4. Deep learning approach

We framed detecting BOS from CT scans as a classification task. We trained a DNN to take a CT scan as input and return an estimate of the log-likelihood that the patient suffers from BOS. DNNs are a well-studied technique for image classification ^21^ but generally need to be trained on large volumes of annotated data to attain high performance. The modest number of patients in our data set makes training prone to overfitting – a phenomenon where the DNN learns to classify training scans with high accuracy but fails to classify examples not seen during training. We employed several techniques to mitigate this adverse effect. First, we used the late-fusion architecture ^22^, which relies on a two-dimensional DNN to process scan slices independently from one another and then fuses the results to produce the disease likelihood. Compared to architectures that rely on three-dimensional representations of the scan, late fusion DNNs have a significantly lower number of parameters, which is crucial for reducing overfitting. Second, we randomly discarded scan slices during training, forcing the DNN to learn recognizing BOS from incomplete scans. This discourages the DNN from classifying training scans using anatomic details or pathologies specific to individual patients, since large portions of the scans are removed at random. Finally, we co-trained the DNN in BOS detection and a novel auxiliary task: temporal precedence prediction of CT scans. Given a random pair of scans of a patient with BOS taken at least six months apart, the auxiliary task is to predict which of the scans was acquired earlier. This allowed us to include CT scans of BOS patients acquired before the patients met BOS clinical criteria in the training of the DNN. These scans would otherwise not be used for training since it is not clear which of them represent signs of the disease and which do not. We provide more details of the architecture of our DNN, the training procedure, and the experiments we performed to validate the design in the supplementary material sections B and C.

## 2. Results

### 2.1. Study participants

All participants were age and sex matched (p-value 0·78 and 0·54, respectively). To compare the lung function between patients of the two groups, we evaluated their relative FEV_1,_ computed as the ratio of the current FEV_1_ to the best FEV_1_ level, defined as the average of two best measurements, after the transplantation, and at least three weeks apart, as recommended by the current clinical practice guidelines ^20^. While the difference between the results obtained three months after the transplantation was not significant, thirty months after transplantation, several BOS patients showed a marked decrease in FEV_1_ (p-value 0·0037) **(Table 1)**.

**Table 1:**
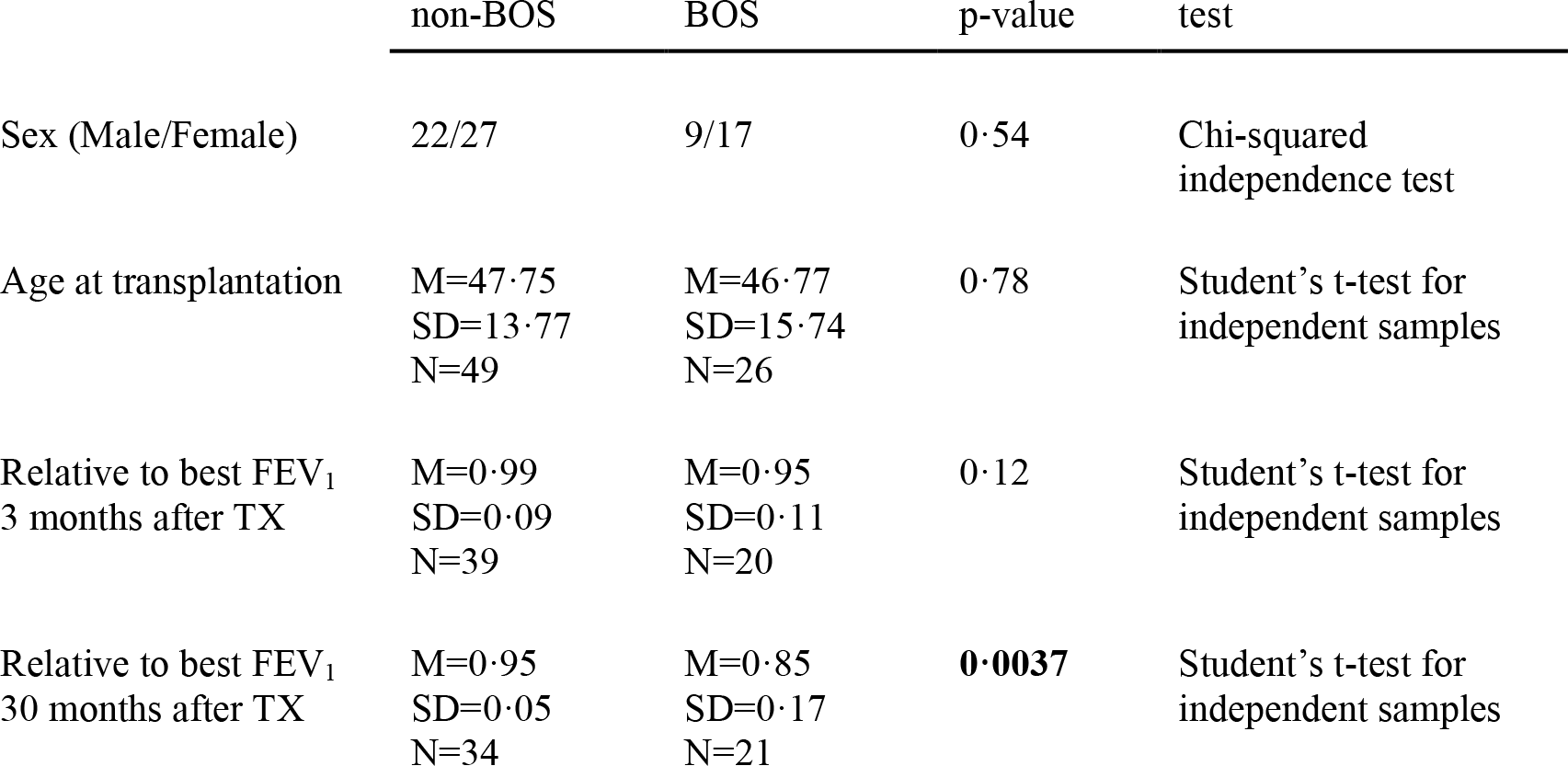
Participant characteristics. Characteristics of patients with or without BOS including sex, age, and FEV1 relative to best FEV1 at 3 or 30 months post lung transplant. M – mean, SD – standard deviation, N – number of patients. TX – transplant. N lower than 26 for BOS patients and 49 for non-BOS patients means that no PFT has been recorded within one month of the query date.

The median time to BOS onset was 3 ·8 years. One thousand days after transplantation, 42% of BOS patients with documented PFT results maintained relative FEV_1_ above 90% of the best level. This fraction diminished to 19% at two thousand days post lung transplant **(Figure 2)**. Total number of patients decreased with time as patients were transferred to other medical centers, underwent a second transplantation, or passed away.

**Figure 2:**
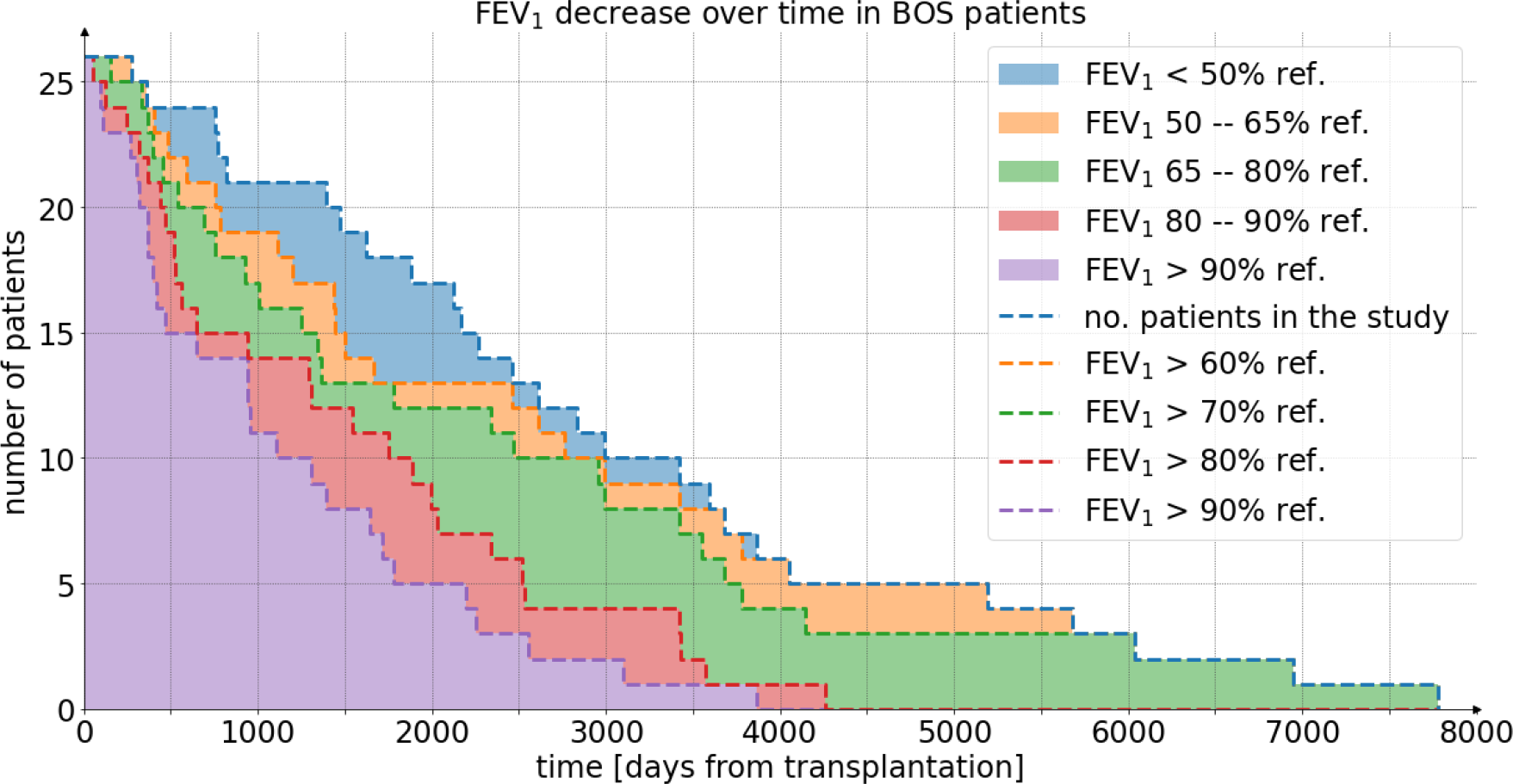
PFT decline after lung transplantation in patients diagnosed with BOS. The height of the bar of each color represents the number of patients in each range of relative FEV_1_. The total height of the colored area corresponds to the total number of patients for whom PFT results were available at the given time.

### 2.2. Detection of BOS

We assessed the performance of our method in distinguishing between CT scans of BOS patients meeting the clinical criteria of BOS stage I or higher and CT scans of patients who were never diagnosed with BOS. The method was able to distinguish patients with BOS from non-BOS with an AUC median of 0·92 varying between 0·87 to 0·95 on individual splits. The aggregated AUC, less affected by the number of patients in individual splits, attained 0·90 (95% CI: 0·840-0·953) (**Figure 3**).

**Figure 3:**
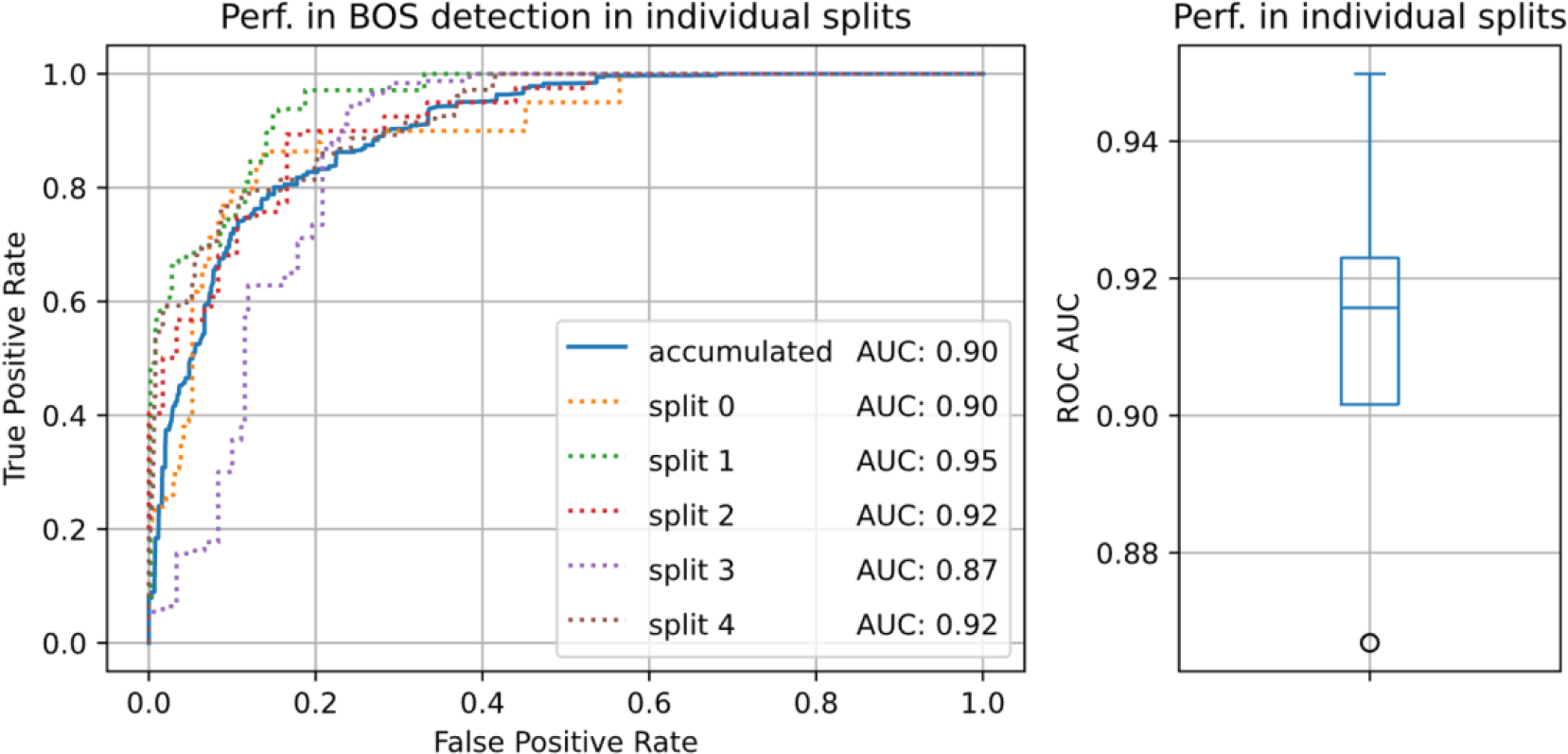
The performance of the deep-learning based method in distinguishing CT scans of BOS from non-BOS patients. Left: weighted ROC curves for individual splits (dotted lines), and the ROC curve resulting from aggregating all splits (continuous blue line); Right: ROC-AUC values of the dotted curves in the left summarized in a box plot.

To understand how the BOS detection accuracy changes as the disease progresses, we tested our method’s ability to distinguish between CT scans of patients within BOS stages I, II, and III, and CT scans of patients who were never diagnosed with BOS. The performance increased with the advancement of BOS: At BOS stage I, the ROC AUC attained 0·88 (CI: 0·798-0·945) and increased to 0·91 (CI: 0·829-0·964) and 0·94 (CI: 0·869-0·989) for stages II and III, respectively. Performance on individual data splits revealed a similar pattern, with a median ROC AUC of 0·89 for stage I, 0·93 for stage II, and 0·98 for BOS stage III (**Figure 4)**.

**Figure 4:**
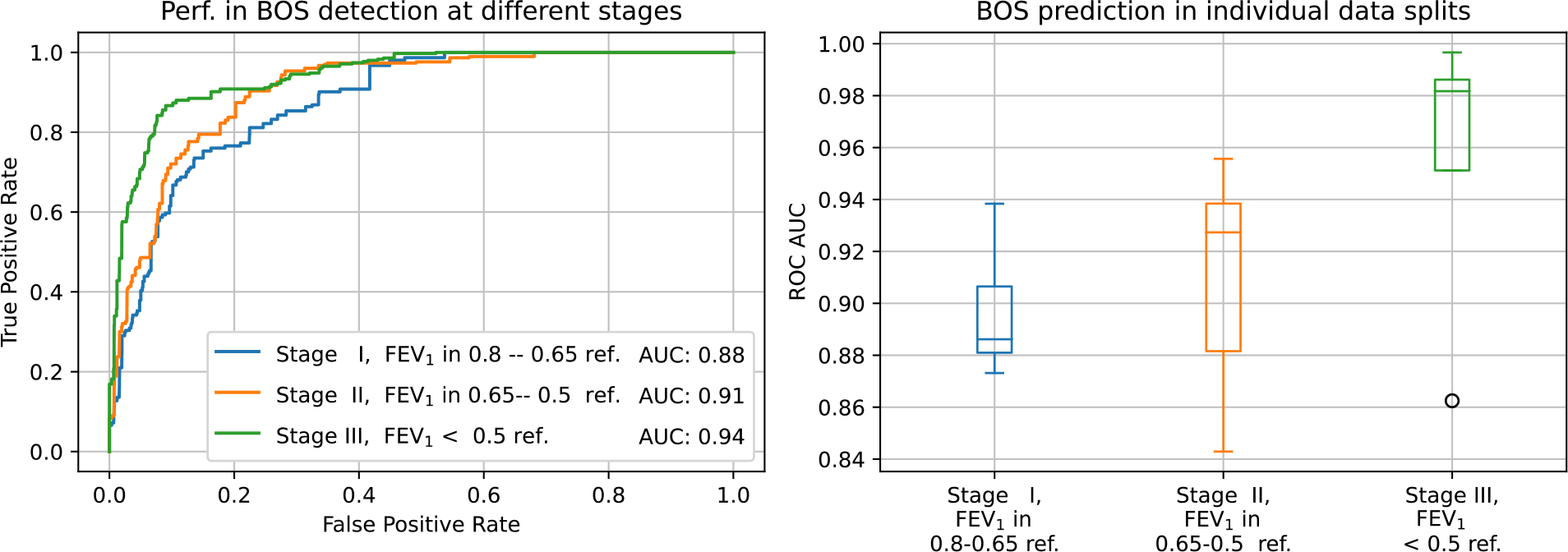
Performance of the method for detecting BOS at different stages of the disease. Left: aggregated weighted ROC curves representing performance of the deep neural network in distinguishing thoracic CT scans of BOS patients at different stages of the disease from scans of patients that were not diagnosed with BOS. Right: ROC-AUC values for individual splits.

### 2.3. Risk prediction for future development of BOS

To verify the capacity of our method to detect patients at risk of developing BOS before they meet the clinical criteria, we evaluated the performance of the network in distinguishing CT scans of BOS patients before the diagnosis with FEV_1_ between 80% and 90% of best FEV_1_ from CT scans of non-BOS patients with FEV_1_ in the same range. The network attained an aggregated ROC-AUC of 0·87 (CI: 0·735-0·974), and between 0·75 and 1·0 on individual splits. The variance of this result can be attributed to the low number of CT scans in the considered FEV_1_ range (11 BOS and 20 non-BOS patients). To contrast these results with a yet more challenging scenario, we also evaluated our method in distinguishing scans of BOS and non-BOS patients with FEV_1_ in the range of 90–100% of the baseline, where we expect to see much less difference between the patients with and without BOS. At this range of FEV_1_ the aggregated AUC decreased to 0·61 (CI: 0·412-0·792) (**Figure 5**). This points to either the absence of disease at this stage, or that the abnormalities are too subtle for our method to detect.

**Figure 5:**
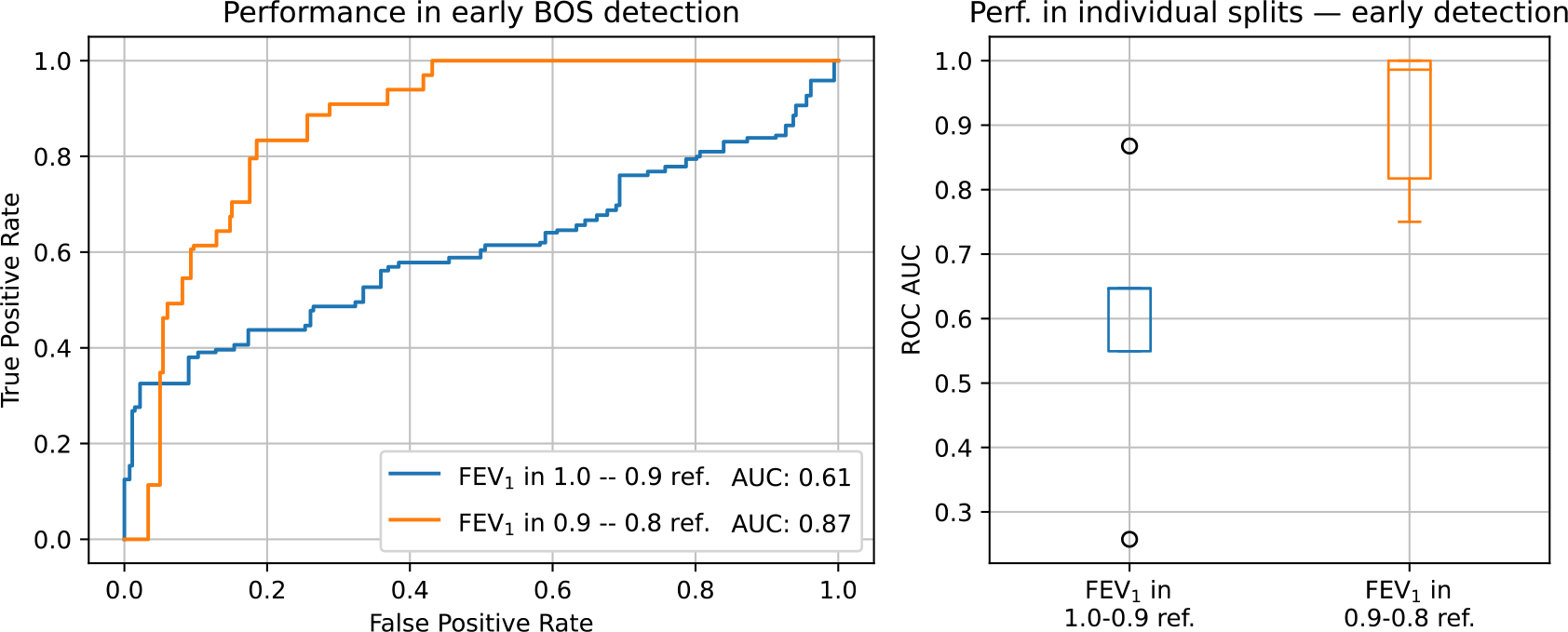
Performance of DNN in the prediction of BOS. Deep network prediction of future onset BOS in patients with relative FEV_1_ in the range of 90–80%, (orange), and 100– 90% (blue). Left: aggregated ROC curves. Right: ROC AUC computed on individual splits.

### 2.4. Detection based on high- and standard-resolution scans

Radiological changes associated with advanced BOS have traditionally been elucidated using high-resolution CT scans acquired during the exhalation phase, necessitating higher radiation doses and specialized protocols. In addition to using CT scans at any stage of respiration as input (see section 1.2), we probed the impact of image resolution—both high and low—on our method’s performance. The dataset for this study comprised 81 high-resolution CT scans from 22 patients with BOS, taken post-disease onset, and 216 such scans from 49 patients without BOS. Additionally, 95 standard-resolution scans were available for 26 patients with BOS, and 176 such scans for 48 patients without BOS. Certain scans existed in both standard- and high-resolution formats, thus the numbers reported here deviate from those in section 2.1. Both types of scans were included in training and evaluating the method to produce the results reported in sections 2.2 and 2.3. To verify if higher scan resolution makes it easier for the DNN to detect BOS, we tested it separately on high- and standard-resolution scans. Remarkably, the method exhibited comparable performance in both scan types, achieving an aggregated ROC-AUC of 0.90 for high-resolution scans (the blue-continuous plot in **Figure 6**) and 0.89 for standard-resolution scans (the blue-dashed plot in **Figure 6**). To confirm this observation, we re-trained the DNN exclusively on high resolution scans. When tested on high-resolution scans, the re-trained DNN demonstrated performance comparable with the one trained on both scan types, attaining a ROC-AU C of 0·87 (the orange-continuous plot in **Figure 6**). But when tested on standard-resolution scans, the re-trained DNN performed remarkably worse, reaching a ROC-AUC of 0·81 (the orange-dashed plot in **Figure 6**), as expected. In summary, limiting training or testing to high-resolution scans did not result in improved performance.

**Figure 6:**
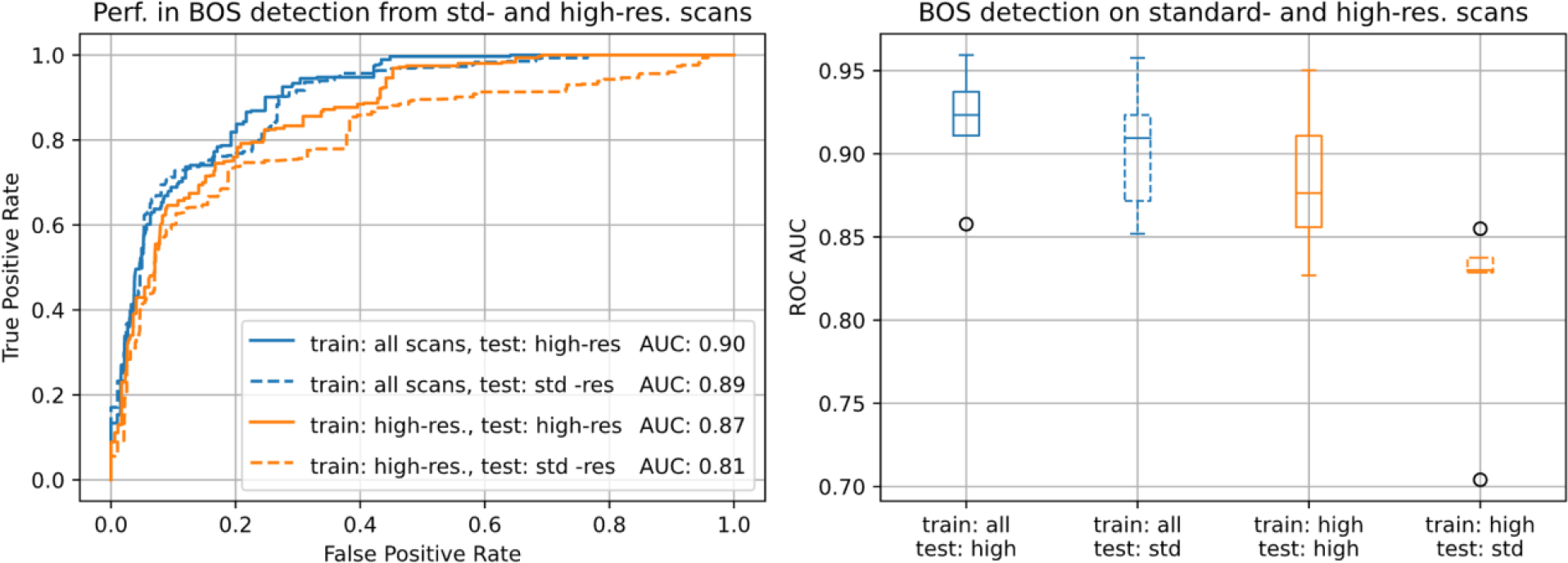
The performance of DNN following training with low or high resolution CTs. Our deep network was trained on standard- and high-resolution scans and could detect BOS in both types of scans (blue). Re-training the network exclusively on high-resolution scans gives on-par performance for high-resolution scans, but significantly lower performance for standard-resolution scans (orange). Left: aggregated ROC curves. Right: AUC attained in individual splits.

### 2.5. Verification of our deep learning approach

To demonstrate the contribution of each of the deep learning techniques to performance, we re-trained the DNN with each of the techniques switched off and evaluated it in distinguishing BOS from non-BOS in the same way as reported in section 2.2. We present the results in **Supplementary Table 1** and **Supplementary Figures 2, 3, and 4**. The results confirm that each of the techniques contributes to the result. Notably, disabling all the techniques, which reduced our method to a standard deep learning approach, dropped the performance to 0·74 (CI: 0·629-0·831) ROC-AUC **(Supplementary Table 1 and Figure 2)**. This illustrates the importance of mitigating overfitting when deploying DNNs in small-scale studies.

### 2.6. Generating explanations for the deep neural network’s decisions

To delineate the lung regions instrumental in the DNN’s diagnostic process, we employed Guided Grad-CAM^23^, a technique to produce masks that highlight the image areas important to the network’s decision-making. In essence, Guided Grad-CAM utilizes the DNN’s gradient to accentuate regions in the image whose alteration would bolster the decision. An example of this explanation map is provided in Figure 7. Remarkably, the marking consistently manifests in hyperlucent areas, compatible with either air-trapping or bronchiectasis. The same focal lesions are marked in both high- and low-resolution scans. Therefore, these areas may represent focal regions utilized by the DNN in its diagnostic assessment.

**Figure 7:**
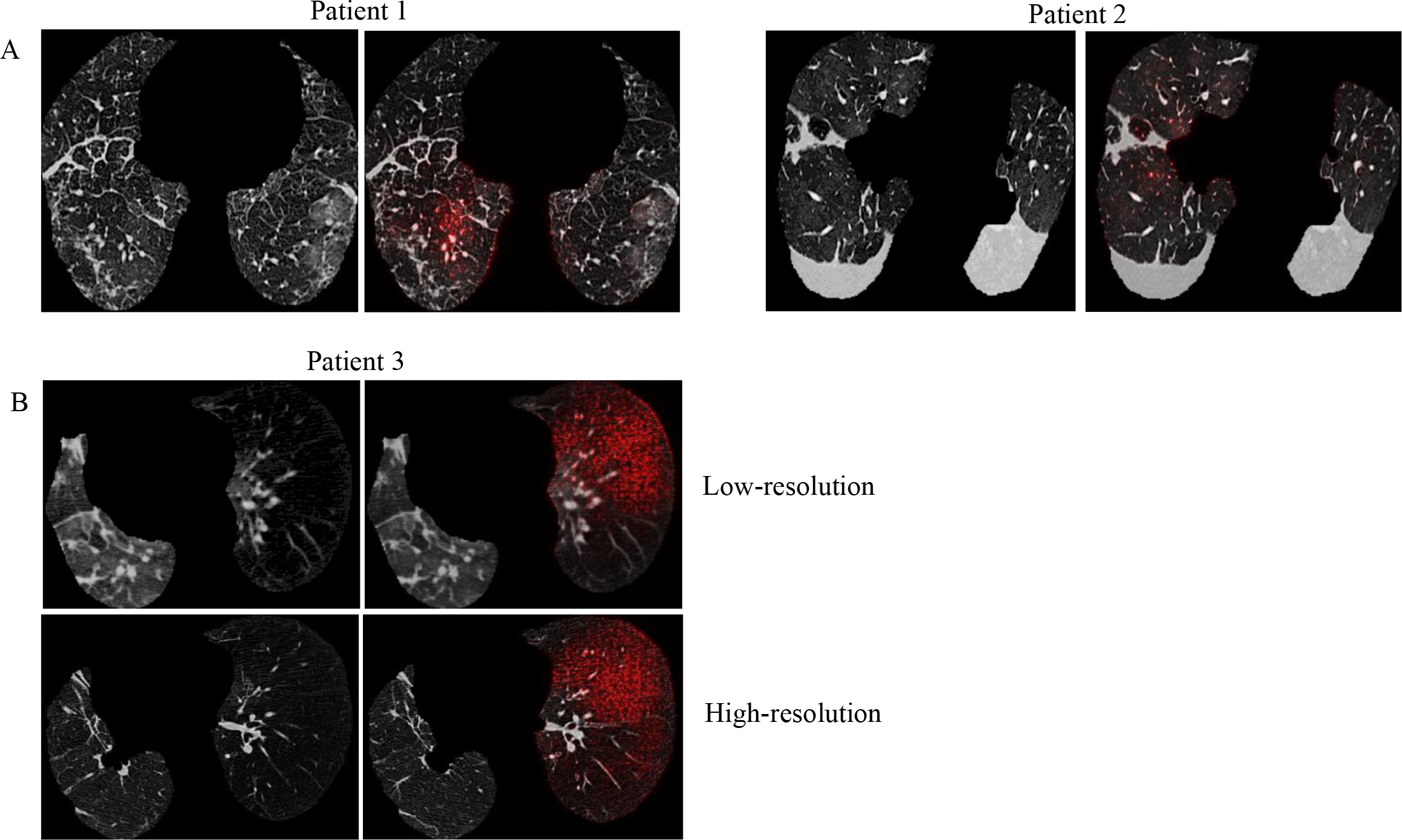
A. visual explanation of the DNN’s decision for two representative BOS patients. Left: a CT slice; Right: the same slice with the attention map obtained using Grad-CAM algorithm. B. DNN interpretation on low- and high-resolution CTs obtained at the same radiology visit.

## 3. Discussion

While DNNs have shown remarkable success in detecting lung diseases such as Covid-19, their application to BOS detection has to date remained unexplored. Importantly, our method improves diagnoses of BOS even in the absence of dedicated CT scans for BOS diagnosis, for example, during full inspiration, excluding the need for special CT protocols during exhalation. Additionally, it can be applied to standard-resolution CT scans, mitigating radiation concerns, and expanding accessibility.

Previous methods of CT scan analysis for BOS detection rely on pre-defined procedures to identify patterns in the lung and cannot benefit from signs of BOS not captured by these procedures.^10,11,12,13,14,15,16,17,18^ However, designing procedures to extract all pertinent information from the scans may not be feasible. Prior research suggests that BOS can manifest itself in non-obvious ways, including by alterations to the volume and surface of the airways, modifications of the volume of pulmonary vasculature ^24,17^, and changes in the density of microvasculature^25,26^, all of which may affect the appearance of the scans. This prompted us to take a different approach and use a DNN that does not rely on fixed pattern identification procedures but instead learns to extract pertinent information directly from the scans. When benchmarked in distinguishing scans of patients with and without BOS, our DNN attains an area under the receiver operating characteristic (ROC-AUC) of 0·90 (CI: 0·840-0·953).

Interestingly, we could train the DNN to detect BOS in non-dedicated, standard-resolution CT scans at any stage of inspiration. The high performance of the resulting BOS detector suggests that current diagnostic procedures might not fully benefit from the information contained in CT scans, and that DNNs can help unlock currently unused information.

The ability to identify BOS in routinely conducted CT scans, without the necessity for high-resolution or paired inspiratory-expiratory scans, implies that our deep-learning-based BOS marker can be computed at reduced expense and without exposing patients to additional procedural risks. This stands in contrast to invasive procedures like surgical lung biopsies, which, although informative, are not routinely used due to the risks of adverse events. The integration of data from various non-invasive modalities such as CT scans and PFTs has the potential to create a more comprehensive diagnostic profile of BOS, improving diagnostic accuracy and contributing to a more nuanced understanding of disease progression.

Moreover, our method shows the promise of early BOS detection, reaching a ROC-AUC of 0·87 (CI: 0·735-0·974) in patients whose lung function decreased but remains above BOS clinical criteria. These results render DNNs a promising avenue for improving BOS diagnosis and enabling early detection, crucial for improving patient outcome. This result is well aligned with previous reports of predicting BOS onset based on airflow simulations and computations of airway volume in lung models reconstructed from CT scans ^24^. In contrast to this approach, our DNN can be applied directly to CT scans, without the need of constructing airway models. Further research is needed to establish a standard of early management, such as azithromycin or extracorporeal photopheresis^27,28^, that could be administrated to patients with elevated risk of developing BOS, identified by a prognostic method, like our DNN.

The effectiveness of the DNN in our study is significantly influenced by strategies employed to counter overfitting, notably our innovative co-training configuration: A DNN trained without these techniques attains a ROC-AUC of 0·74 (CI: 0·629-0·831), which represents a large performance drop from the 0·90 (CI: 0·840-0·953) ROC-AUC attained by our method. Anticipatedly, these techniques will diminish in significance with the availability of a sufficiently extensive training dataset. However, financial constraints often restrict the size of patient cohorts accessible for the initial assessment of novel technologies in diagnosing diseases with low incidence. In this context, our method could offer valuable benefits to future studies facing similar limitations.

To conclude, we described a novel method for detecting BOS through cutting-edge deep learning applied to CT scans. Our approach holds clinical relevance in lung transplantation, thanks to the capacity to detect BOS early, pivotal in enabling timely interventions and improving patient care.

## Data Availability

All data produced in the present study are available upon reasonable request to the authors

## 4. Acknowledgement

This work was supported by the FWF Austrian Science Fund Lise Meitner grant number M3374 to MK, and by SNSF grants CRSK-3_190526 and 310030_204938 to SJR. The funders of the study had no role in study design, data analysis, data interpretation, or writing the manuscript.

