## supplementary material for "Harnessing Deep Learning to Detect Bronchiolitis Obliterans Syndrome from Chest CT"

### Supplementary Material: Harnessing Deep Learning to Detect Bronchiolitis Obliterans Syndrome from Chest CT Scans

In this supplementary material, we complement the main manuscript with details of our deep-learning-based method and its experimental evaluation. Since our method differs from approaches previously applied to detecting BOS from thoracic CT scans, we first discuss these approaches and the motivation for our method in section A. Next, we describe the details of our method in section B. Finally, we provide additional information about the experimental evaluation and additional experiment results in section C.

#### A Related work: detecting lung diseases from CT scans

To put our work in context, in this section we map the landscape of previous research on automatic detection of lung diseases. We focus on three key areas. First, we review methods for automated BOS detection from CT scans and show that they rely on heavily engineered feature extraction procedures. By contrast, our approach employs deep learning and detects BOS directly from CT scans. We review similar learning methods, previously used in COVID-19 detection, in the second subsection. Since our deep learning approach includes a novel co-training setup, we devote the third part of this section to related techniques.

**Detecting BOS from CT scans** Previous work on computational BOS detection from CT scans is based on hand-crafted 'feature extraction' procedures that extract information pertinent to the disease from the scans. Virtually all these methods follow the same design. First, they assign a 'phenotype' to each voxel of the scan and compute the phenotype frequency, that is, the fraction of the total lung volume taken by each phenotype. In the most popular approach, called Parametric Response Mapping, the phenotype represented by each voxel is recognized by thresholding the intensity of the voxel in a pair of inspiration-expiration scans, aligned using a deformable registration technique.[1–7] Other methods rely on local texture classifiers, pre-trained to imitate radiologists in identifying lung texture types.[8,9] Once the texture type of each voxel has been determined, the phenotype frequency is computed as the ratio of the number of voxels representing the phenotype to the total number of lung voxels. In addition to the phenotype frequency, previously used techniques include measures derived from the histogram of the scan,[10] the volume and surface of individual lung lobes, and aerodynamic characterization of airway models,[4] reconstructed from CT scans using a semi-automatic procedure. Previous studies investigated the predictive value of individual such measures[1,2,6–9] or combined several of them by training an SVM to predict the diagnosis.[3–5] These 'computational BOS markers' were shown to be statistically different when computed for BOS and non-BOS scans,[1–5,9] and scans showing other types of Chronic Lung Allograft Dysfunction,[7–10] formerly called chronic allograft rejection. The best performance in differentiating BOS from non-BOS scans, reported by Sharifi et al.[5] for patients after bone marrow transplantation, reached 0·85 ROC-AUC.

The main disadvantage of the techniques discussed above stems from their reliance on pre-defined procedures to extract information from CT scans. It is very difficult to design feature extraction procedures that would be effective across a range of scan resolutions and for scans taken at any stage of inspiration. We therefore adopt a different approach, based on deep learning, where a deep neural network is trained to detect BOS directly from CT scans. We review previous applications of deep learning to thoracic CT scan classification in the next section.

**Deep learning for detecting lung diseases from CT scans** Deep learning is a machine learning approach that outperforms previous methods of visual recognition by a vast margin.[11] Deep neural networks (DNNs) – deep learning classifiers – are constructed by compositing many linear and nonlinear functions, commonly called layers. The definition of the layers and their composition are referred to as the architecture of the DNN. The first layer of a DNN operates on intensity values of the input image, while the last layer produces the desired output, for example, a log-likelihood of the class of the object shown in the image. The capacity to learn end-to-end, in other words, to learn the entire mapping from a raw image to the likelihood of its class, distinguishes DNNs from previous machine learning techniques, which rely on fixed, pre-defined procedures to extract key information from the image and limit learning to a classifier that maps this information to class likelihood. The end-to-end design gives DNNs more flexibility in learning the decision function and is the main reason for their superior performance.

The first applications of deep learning to thoracic CT scan classification included predicting longevity[12] and detecting COPD.[13] The COVID-19 pandemics, and the COVID-19 detection challenge[14] catalysed research in this direction and led to strong results in COVID-19 detection.[15–23] Unfortunately, the high accuracy of this approach is predicated on the availability of a large volume of training data. Training on small data sets provokes overfitting – a phenomenon in which the DNN attains high accuracy on the training examples, but fails to classify scans not seen during training. Overfitting is the main factor inhibiting the application of deep learning to studies including only modest numbers of patients, which is very often the case with studies focused on diseases of lower incidence rates, like BOS.

A common way to alleviate overfitting is to slightly change the problem setting and train a DNN to segment disease-provoked lesions in a CT scan instead of classifying the scan as representing either a healthy or a diseased lung. By contrast to scan classification, which produces a single disease likelihood for the entire scan, segmentation consists in assigning a lesion likelihood to each voxel of the scan. Training annotations take the form of masks indicating the presence or absence of a lesion in each voxel and guide the DNN to recognize areas of the scan representing the disease. This method proved effective in segmenting lesions provoked by COVID-19[24–28] and Interstitial Lung Diseases.[29,30] Unfortunately, it would be difficult to extend it to BOS, which provokes minute lesions distributed over the lung. Comprehensive annotation of such disease manifestations is virtually impossible, and leaving them unannotated might compromise performance of the DNN. We therefore assumed the more challenging approach, in which the DNN is trained to classify scans, and combined several techniques to address overfitting.

**Co-training DNNs in thoracic CT scan classification and auxiliary tasks** Training a neural network in several tasks simultaneously is a well studied technique to improve its performance.[31] The choice of the auxiliary task is an art in itself: sharing a deep network between multiple tasks makes most sense if the tasks are related, for example, if solving each of the tasks requires extracting the same information from the input data. The auxiliary tasks used for training CT scan classifiers include lung segmentation,[32] segmentation of lung lesions,[33,34] and predicting if a pair of lungs represents the same disease.[18] Producing annotations required to train the network in the auxiliary task is typically too time consuming, so self-supervised tasks, like autoencoding the scan[28] and predicting relative location of patches extracted from the scan,[22] are better suited for most practical scenarios. To our knowledge, we are the first to use predicting temporal precedence between a pair of lung scans as the auxiliary task. It does not require annotating the scans and reflects a simplistic BOS model, in which progression of the disease should be observed between two scans, provided the period between them is sufficiently long.

#### B Our deep learning approach

To deploy deep learning for detecting BOS from CT scans, we frame the problem as a classification scenario,[11] in which a deep neural network (DNN) consumes a CT scan and returns an estimate of the log-likelihood that the patient suffers from BOS. We use the ResNet-18,[35] a proven architecture with low memory requirements, and use the late fusion approach to adapt it to processing three-dimensional scans. We present it in Figure 1. Before we discuss this adaptation in detail, in the next paragraph we introduce the standard training procedure, which serves as a foundation for our method.

**Standard training procedure** We denote the DNN by $f$ and its parameter vector by $\theta$. To adjust $\theta$, the network is trained on a data set $T$ of pairs $(\mathbf{x}, \hat{y})$, where $\mathbf{x}$ is a training scan and $\hat{y} \in \{0, 1\}$ is the ground truth label. $\hat{y} = 1$ represents the presence of BOS and $\hat{y} = 0$, its absence. Training deep networks is formalized as solving

$$\min_{\theta} \sum_{(\mathbf{x},\hat{y}) \in T} L(f(\mathbf{x};\theta), \hat{y}), \tag{1}$$

where the loss function $L$ measures the discrepancy between the prediction $f(\mathbf{x}, \theta)$ and the label $\hat{y}$. We take $L$ to be the binary Cross Entropy, defined as

$$L(f(\mathbf{x};\theta), \hat{y}) = \begin{cases} -\log \frac{e^{f(\mathbf{x};\theta)}}{1+e^{f(\mathbf{x};\theta)}} & \text{if } \hat{y} = 1 \\ -\log \frac{1}{1+e^{f(\mathbf{x};\theta)}} & \text{if } \hat{y} = 0 \end{cases}. \tag{2}$$

The minimization is performed using an iterative, gradient-based method. In practice, due to computational limitations, the gradient of the objective function (1) is estimated on randomly composed batches of the training data as opposed to the entire training set, and the rule for updating $\theta$ employs some form of a momentum mechanism to offset the stochasticity of the gradient.[36] We use the ADAM rule,[37] which normalizes the gradient approximation using estimates of the first- and second-order moments of the distribution of the observed gradients.

**The main challenge** The standard deep learning approach described above works well in scenarios where training data is abundant. Unfortunately, this is not the case with our study. The modest number of patients in our data set makes training prone to overfitting – a phenomenon where the network learns to classify training scans with high accuracy but fails to classify examples not seen during training. In case of classifying CT scans, the risk of overfitting is aggravated by the fact that each scan is rich in information unrelated to BOS. In consequence, it is easy for the network to classify the training scans using this information, as opposed to lesions provoked by the disease. A common way to circumvent this problem is by training the network to identify areas of the scan affected by the disease.[24–29] However, this requires annotating all disease-affected regions in each training scan. In case of BOS, which often provokes minute lesions dispersed over the lung, such annotation is particularly challenging. We therefore forego this approach and simply train our network to predict whether a patient is affected by BOS or not. To mitigate overfitting, we employ a compact late-fusion architecture, drop entire scan slices during training, and propose a novel co-training setup. In the remaining part of this section, we describe each of these methods in detail.

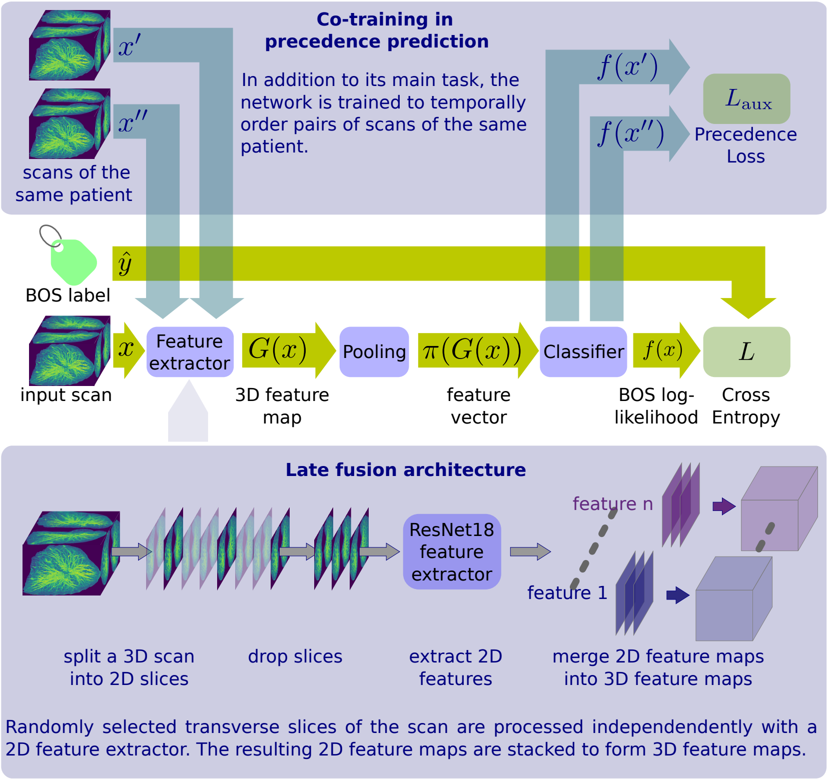

Figure 1: The diagram of our training setup. The pipeline in the middle represents the standard training procedure. The blue boxes illustrate adaptations for BOS detection.

**Late fusion architecture** We give our DNN the late fusion architecture[13,14,21,38] which relies on a 2D feature extractor to process scan slices independently from one another, then fuses the resulting 2D feature maps with a voting or pooling operation, and produces the disease likelihood with a fully-connected classification network. We illustrate it in Figure 1. The late fusion architecture has several advantages over architectures that heavily rely on 3D representations: it features a lower number of parameters, which reduces the risk of overfitting, requires less computation, and facilitates adopting a pre-trained DNN as the feature extractor. Fine-tunning a DNN pre-trained on a large data set in a task different from the target one is an effective way to improve performance and speed up training convergence,[39,40] but is often difficult to implement for 3D architectures, because pre-trained 3D DNNs are not commonly available. By contrast, DNNs pre-trained on large sets of 2D images are publicly available, and the late fusion approach enables adopting them as the backbone of the architecture. We now describe the application of this approach to the construction of our DNN for BOS detection.

The core of our DNN is the convolutional feature extractor of ResNet18,[35] obtained by removing the topmost linear layer and the pooling layer from the full ResNet18 architecture. We take the feature extractor of a ResNet18 pre-trained on ImageNet,[41] a data set comprising more than one million images, and denote this feature extractor by $g$. Given a CT scan $\mathbf{x}$, with $D$ transverse slices of height $H$ and width $W$, we apply $g$ to each slice of the scan. For each slice of the scan, this results in a two-dimensional feature map $g(\mathbf{x}_j)$, where $\mathbf{x}_j$ denotes the $j$-th slice of the scan. $g(\mathbf{x}_j)$ is a table of height $h = H/32$ and width $w = W/32$ containing feature vectors with 512 components. We denote the $i$-th component of the feature vector at position $(k, l)$ of the 2D feature map by $g(\mathbf{x}_j)[i, k, l]$. We stack the 2D feature maps $g(\mathbf{x}_j)$ to obtain a 3D feature map $G(\mathbf{x})$ of depth $D$, height $h$, and width $w$. The $i$-th component of the feature vector at position $(j, k, l)$ of the 3D feature map is defined as $G(\mathbf{x})[i, j, k, l] = g(\mathbf{x}_j)[i, k, l]$.

The 3D feature map $G(\mathbf{x})$ is reduced to a single vector with the max-pooling operation $\pi$. The $i$-th component of the vector resulting from pooling is defined as

$$\pi(G(\mathbf{x}))[i] = \max_{j,k,l} G(\mathbf{x})[i, j, k, l], \tag{3}$$

where $j$, $k$, and $l$ index the depth, height, and width of the feature map, respectively. This vector is then forward-propagated through a linear classification layer with one output variable to produce the log-likelihood of BOS. We denote the vector of weights of the linear layer by $\mathbf{w}$, and its scalar bias by $b$, and define our full neural network as

$$f(\mathbf{x}) = \mathbf{w}^\intercal \pi(G(\mathbf{x})) + b. \tag{4}$$

**Randomly discarding scan slices** Following previous work,[13] during training we process only eight randomly selected slices of each training scan, and discard the remaining ones. This forces the DNN to detect manifestations of the disease in multiple slices of the scan, because individual slices are discarded at random. It also drastically limits the information overlap between two presentations of the same scan. Finally, it reduces the computation required to process each training scan.

**Co-training with precedence prediction** When training the network to detect BOS, we use the scans of patients diagnosed with BOS, taken after the diagnosis, and the scans of the patients that received a transplantation but were not diagnosed with BOS. We refrain from using the scans of BOS patients taken before they met BOS clinical criteria. Since these scans might contain no hint of the disease, using them as negative training examples could compromise the DNN's capacity to diagnose early BOS. Nevertheless, these scans represent an opportunity to train the network to differentiate between early and late stages of the disease in the same patient. To benefit from this opportunity, we co-train the network in BOS detection and an auxiliary task: given a random pair of scans of the same patient, taken at least six months apart, predict which one of them was acquired earlier. We perform this training exclusively for patients that ultimately developed BOS.

Given a pair of scans $(\mathbf{x}', \mathbf{x}'')$ of the same patient, where $\mathbf{x}'$ has been acquired before $\mathbf{x}''$, we compute the likelihood that $\mathbf{x}''$ has been acquired later using the softmax function: $\frac{e^{f(\mathbf{x}'')}}{e^{f(\mathbf{x}')}+e^{f(\mathbf{x}'')}}$. We use this formulation to train the network in the auxiliary task with the cross entropy loss

$$L_{\text{aux}}(f(\mathbf{x}'), f(\mathbf{x}'')) = -\log \frac{e^{f(\mathbf{x}'')}}{e^{f(\mathbf{x}')} + e^{f(\mathbf{x}'')}}. \tag{5}$$

To formalize the complete co-training objective, we denote the set of all possible pairs of training scans of the same patient taken at least six months apart by $T_{\text{aux}}$ and solve

$$\min_{\theta} \sum_{(\mathbf{x},\hat{y})\in T} L(f(\mathbf{x};\theta), \hat{y}) + \alpha \sum_{(\mathbf{x}',\mathbf{x}'')\in T_{\text{aux}}} L_{\text{aux}}(f(\mathbf{x}'), f(\mathbf{x}'')) \tag{6}$$

where $L$ is the cross-entropy loss for BOS classification and $\alpha$ balances the two loss terms. We set $\alpha = 1$. This formulation not only lets us employ the 58 scans of 22 BOS patients taken before they met the clinical criteria for training, but also encourages the network to vary the predictions for scans of the same patients, discouraging it from overfitting to anatomic details of individual patients. As shown by the results of our experiments, it holds a contribution to the performance of our BOS classifier.

**Implementation details** We pre-processed the scans by cropping them tightly around lung masks, obtained by lungmask,[42] a publicly available deep network trained to segment the lung in CT scans. Scan intensity was clipped to between −1000 and 600 Hounsfield Units, and re-mapped to the interval $[-1; 1]$. Subsequently, we rescaled the images to match the pixel size of 0·5 mm and the inter-slice spacing to 1·0 mm.

Training the DNN was performed using the PyTorch library. The batch size was set to 10 for both the BOS classification task and the temporal precedence recognition task. We took random crops of the scan of size 512 by 284 pixels in the transverse plane and kept randomly selected eight slices of the resulting 3D volume. Data augmentation consisted in random flips of the scan in the transverse plane. Training was performed using the ADAM update rule. We set the learning rate of 1e-4 for the first 50 thousand iterations, and then divided it by two every five thousand iterations until 65 thousand iterations.

Since our patient cohort is too small to split out a separate validation set, we did not perform model selection. To avoid biasing the reults to the test sets, we simply kept the network after the last parameter update as the final result of our training procedure.

#### C Experimental evaluation

This section complements the results and methodology presented in the paper with additional details and extended experimental evaluation. In subsection C.1, we define the used performance metrics. Subsection C.2 is devoted to demonstrating the contribution of the individual design decisions behind our deep learning setup to performance. Subsection C.3 offers additional insight into the performance of our DNN.

#### C.1 Performance metrics

We used performance metrics based on the receiver operator characteristic (ROC), a plot of the true positive rate (TPR) versus the false positive rate (FPR), computed by varying the threshold $\tau$ applied to the output of the deep network $f$. Given a set of "positive" scans $P$ and a set of "negative" scans $N$, FPR and TPR can be expressed as functions of $\tau$:

$$\text{FPR}(\tau) = \frac{\sum_{x\in N}[f(x) > \tau]}{|N|} \quad \text{and} \quad \text{TPR}(\tau) = \frac{\sum_{x\in P}[f(x) > \tau]}{|P|}, \tag{7}$$

where $[\cdot]$ equals one when the logical expression inside the bracket is true and zero otherwise. ROC is the parametric curve $(\text{FPR}(\tau), \text{TPR}(\tau))$. The area under this curve (AUC) serves as our performance metric. ROC-AUC ranges between 0 and 1, with 0·5 corresponding to a random prediction, and 1·0 to a perfect one.

Some patients presented to the radiology service more times than others and some visits resulted in more than one copy of the acquired scan, often reconstructed in different resolutions or with different kernels. We systematically included all the copies that satisfied the inclusion criteria presented in the paper. To prevent biasing the performance estimate towards the patients that received more scans, or towards scans with a larger number of copies, we equalized the influence of each patient and each scan by weighting the predictions when computing the AUC. We defined the weight applied to scan $x$ by $w_x = 1/n_{\text{visits}}(x) 1/n_{\text{copies}}(x)$, where $n_{\text{visists}}(x)$ is the number of times the patient presented to radiology, and $n_{\text{copies}}(x)$ is the number of copies of scan $x$. The re-weighted TPR and FPR were computed as

$$\text{FPR}^w(\tau) = \frac{\sum_{x\in N}[f(x) > \tau]w_x}{\sum_{x\in N} w_x} \quad \text{and} \quad \text{TPR}^w(\tau) = \frac{\sum_{x\in P}[f(x) > \tau]w_x}{\sum_{x\in P} w_x}, \tag{8}$$

and the weighted ROC was defined as the parametric curve $(\text{FPR}^w(\tau), \text{TPR}^w(\tau))$.

We used a five-way data split; Each experiment consisted in training and testing the DNN five times, each time with one split of the data kept for testing and the remaining four splits used as the training set. This resulted in five AUC values, which we report in the form of box plots. To summarize the performance in one number, we computed a single ROC curve for test predictions of all five splits, and evaluated the corresponding AUC. We call this metric ‘aggregated AUC’. Unlike simply averaging the AUC values for different splits, the aggregated AUC provides a conservative performance estimate, because each point in the ‘aggregated’ ROC curve results from applying the same threshold to predictions produced by five independently trained neural networks, which likely have different optimal thresholds.

**Confidence intervals** We computed the 95% confidence intervals using hierarchical case resampling,[43] to preserve the structure in the data, where the levels of the hierarchy are: patients, scans, and scan copies. In short, the sampling procedure comprised: first sampling the number of patients equal to the size of cohort in our study, then, for each selected patient, sampling a number of scans equal to the number of scans of the selected patient in our study, and, finally, for each selected scan, sampling a number of scan copies equal to the number of copies of the selected scan included in the study. Thie aggregated ROC-AUC was then computed in the manner described above. We used 10 thousand experiments to estimate the confidence intervals. This approach results in much wider confidence intervals than treating the scan independently, but respects

Table 1: The performance (ROC-AUC) of our DNN, trained once with each key component turned off, in differentiating the scans of patients with BOS from the scans of patients never diagnosed with BOS.

| | split 1 | split 2 | split 3 | split 4 | split 5 | aggregated |
|---|---|---|---|---|---|---|
| full method | 0·90 | 0·95 | 0·92 | 0·87 | 0·92 | 0·90 |
| no co-training | 0·83 | 0·94 | 0·86 | 0·86 | 0·90 | 0·85 |
| no slice dropping | 0·90 | 0·82 | 0·84 | 0·87 | 0·85 | 0·85 |
| 3D DNN | 0·92 | 0·73 | 0·55 | 0·87 | 0·62 | 0·74 |

no co-training – the DNN trained without our co-training method
no dropping slices – the DNN trained without discarding slices
3D DNN – a 3D ResNet, trained without co-training or slice dropping

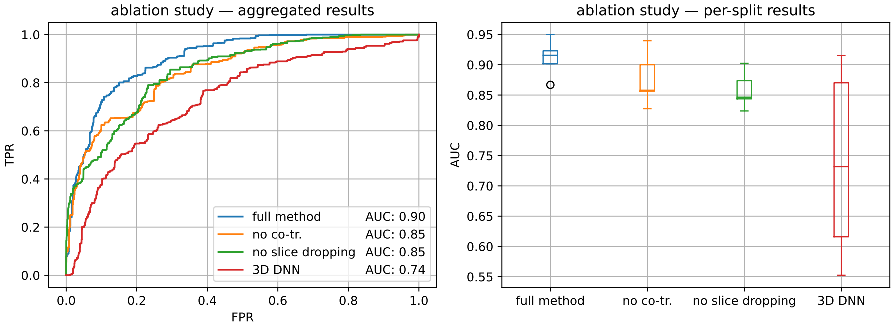

Figure 2: Switching off each of the techniques to limit overfitting compromises the performance of our DNN in distinguishing scans of patients diagnosed with BOS from scans of patients without BOS. Left: aggregated ROC curves illustrating the performance of the DNN trained without individual the techniques. Right: box-plots illustrating the performance of the DNNs on individual splits.
no co-tr. – the DNN trained without our co-training method
no dropping slices – the DNN trained on stacks of 32 consecutive scan slices
3D DNN – a 3D ResNet, trained without co-training or slice dropping

the dependence among multiple scans of the same patient and preserves the number of scans acquired for each patient. Notably, due to the hierarchical structure of the data, the resulting intervals need not be symmetric around the estimated most likely ROC-AUC and their width differs between experiments, due to the differences in the numbers of included patients and their scans.

#### C.2 How elements of our method contribute to performance

To evaluate the design decisions behind our approach, we trained the DNN to differentiate scans of patients with BOS from scans of patients never diagnosed with BOS multiple times, each time with one of the key components of the method, described in section B, switched off. The results are presented in Figure 2 and Table 1. We discuss the evaluated DNN configurations and their performance below.

**Co-training** First, we removed co-training and restricted training to the cross entropy loss for BOS detection (1). As can be seen in the left part of Figure 2, the ROC curve corresponding to the DNN trained without co-training, plotted in orange, is dominated

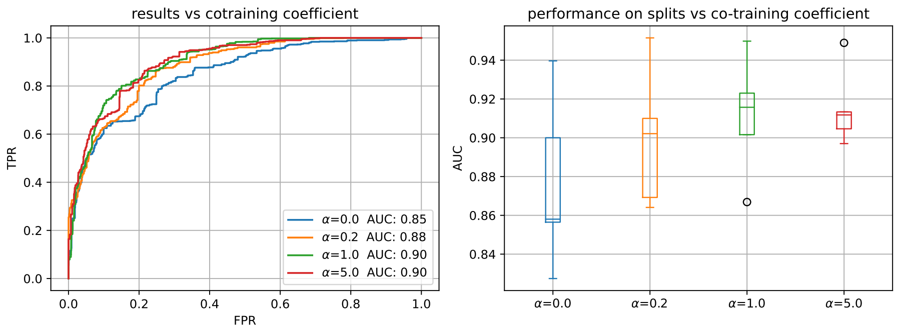

Figure 3: The performance of our DNN versus the co-training coefficient $\alpha$. Left: aggregated ROC curves illustrating the performance of DNNs trained with different $\alpha$. Right: a box-plot illustrating the performance of the DNNs on individual splits.

by the curve attained by training the DNN in the full setup. The numerical results in Table 1 show that the DNN trained in the simplified setup, without co-training, is outperformed by the DNN trained in the full setup on each data split. Removing co-training decreases the aggregated ROC-AUC from 0·90 to 0·85.

To further investigate the influence of co-training on performance, we trained our DNN multiple times with different values of the co-training coefficient, denoted by $\alpha$ in equation (6). We present the results in Figure 3. The aggregated ROC-AUC increased when increasing $\alpha$ from 0 to 1. This confirms the contribution of co-training to performance of our DNN. Increasing $\alpha$ further does not result in any performance gain.

**Dropping scan slices during training** To verify the effect of randomly discarding training scan slices on performance, we trained the DNN on random crops of training scans comprising 32 consecutive slices. Other details of the training setup were kept unchanged. The results, presented in green in Figure 2, show that not discarding scan slices provokes a performance drop from 0·90 to 0·85 aggregated ROC-AUC. Comparing the first and the third row of Table 1 reveals that the DNN trained in the full setup outperforms the one trained without slice dropping on three data splits, and matches its performance on the two remaining ones.

To further confirm the influence of discarding the slices of training scans on performance, we re-trained the DNN with different numbers of retained slices. As can be seen in Figure 4, retaining too many slices defeats the purpose of discarding slices. Similarly, discarding too many slices affects performance, likely due to increased chances of not observing signs of the disease in the slices that were retained.

**3D architecture** To verify the joint contribution of all the design decisions to performance, we trained a 3D DNN, without the late fusion architecture, dropping slices, or co-training. The 3D DNN was constructed by substituting the feature extractor, denoted $G$ in equation 4 and based on applying a 2D ResNet18 to slices of the scan, with a 3D version of the ResNet18 feature extractor.[44] As shown in Figure 2 and Table 1, this approach attained a ROC-AUC of 0·74, a 0·16 drop from the ROC-AUC of 0·90, attained by our method.

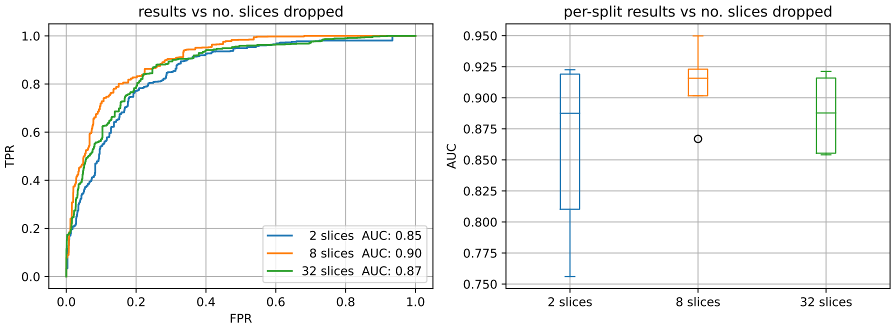

Figure 4: The performance of our DNN versus the number of slices retained for every training scan. Left: aggregated ROC curves illustrating the performance of DNNs trained with different numbers of retained scan slices. Right: a box-plot illustrating the performance of the DNNs on individual splits.

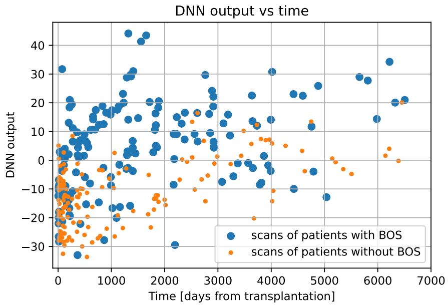

Figure 5: BOS log-likelihood produced by our DNN for scans of patients with and without BOS as a function of time from transplantation.

##### C.3 Additional results

In this section, we investigate the BOS likelihood predicted by our DNN as a function of time and patient's $FEV_1$. Additionally, we motivate future work to focus on distinguishing between BOS and confounding diseases.

**DNN output over time** In Figure 5, we show the output of our DNN as a function of time between the scan and the transplantation. Similarly, in Figure 6, we show the output of our DNN, for patients with BOS, as a function of time between the BOS diagnosis and the scan. The DNN was trained to distinguish between scans of BOS patients, taken after the diagnosis, and scans of patients never diagnosed with BOS, as described in the main manuscript. Like before, we take the diagnosis date to be the date of the first PFT in which the measured $FEV_1$ fell below 80% of the best level and remained on that level on subsequent tests or decreased further. As expected, the predicted BOS log-likelihood increases with time, and a steep transition from low to high likelihood can be observed in Figure 6 near the diagnosis date.

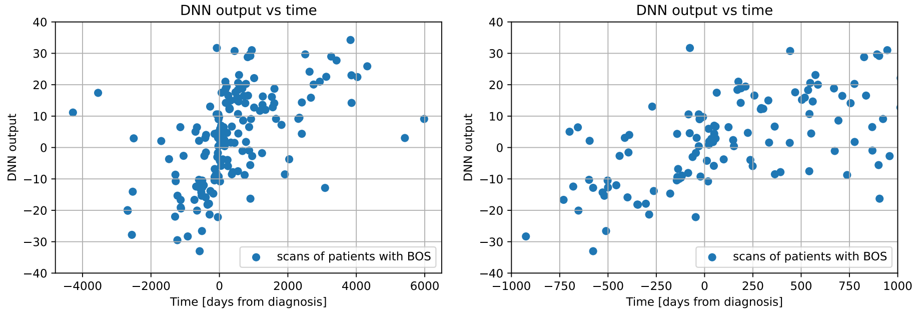

Figure 6: The BOS log-likelihood output by our DNN for CT scans of patients diagnosed with BOS as a function of days passed from BOS diagnosis. Left: full plot, right: with the time axis limited to ±1000 days.

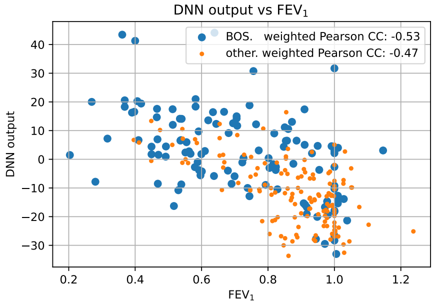

Figure 7: The BOS log-likelihood produced by our DNN for scans of patients with and without BOS as a function of the $FEV_1$ measured within 30 days from the scan date. The predicted BOS likelihood is anti-correlated with $FEV_1$.

**DNN output with respect to patient's $FEV_1$** In Figure 7, we present a scatter plot of BOS log-likelihoods produced by our DNN with respect to patient's $FEV_1$, for patients with and without BOS. As can be seen in the plot, the output of our neural network is correlated with $FEV_1$, attaining the weighted Pearson correlation coefficient of -0·52 for patients with BOS and -0·45 for patients not affected by this disease.

**Distinguishing scans showing BOS from scans of patients without BOS but with $FEV_1$ below 80% of the best value** Figure 5 shows that the BOS likelihood predicted for the scans taken in the first two thousand days after the transplantation behaves as expected: for BOS patients the number of scans assigned an elevated BOS likelihood grows with time, while for patients who were not diagnosed with BOS the predicted BOS log-likelihood remains below zero. However, scans of patients without BOS taken later than two thousand days after the transplantation seem to appear more confusing to the DNN: some of them are assigned elevated BOS likelihood. A similar effect can be observed in Figure 7: some scans of patients without BOS, taken at $FEV_1$ below 70% of the best value, are assigned elevated BOS likelihood. This may be caused by the presence of confounding diseases, including Chronic Lung Allograft Dysfunction of types other than BOS. Patients with such diseases were not specifically excluded from

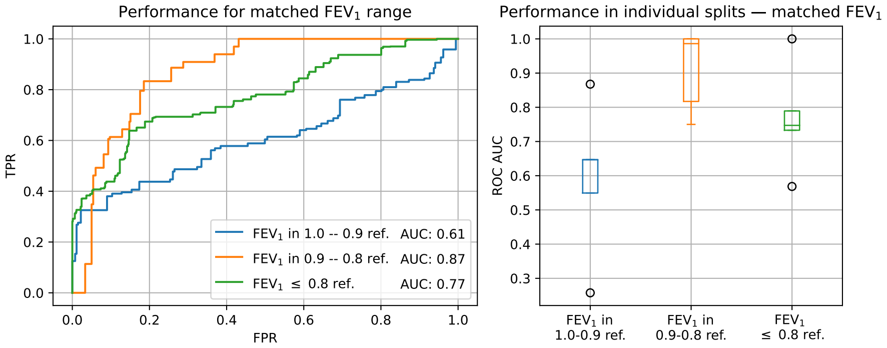

Figure 8: Results of our DNN in distinguishing scans of patients with and without BOS, taken when patients' $FEV_1$ was in the same range of $[1{\cdot}0, 0{\cdot}9]$, $[0{\cdot}9, 0{\cdot}8]$ and $[0{\cdot}8, 0{\cdot}0]$, respectively, for both groups of patients.

the study. As a result, our data set contains 35 scans of 13 patients without BOS, taken when patient's $FEV_1$ was below 80% of the best value.

To investigate this further, we evaluated performance of the DNN in distinguishing between scans of patients with and without BOS, taken at matched $FEV_1$ intervals. As shown in Figure 8, for $FEV_1$ below 80% of patient's best value, the performance attains 0·74 ROC-AUC, which represents a marked decrease from 0·90 ROC-AUC that we reported for distinguishing between the scans of patients with BOS and *all* scans of patients without BOS, irrespectively of their $FEV_1$. The lower performance of the DNN in distinguishing between scans of patients with BOS and patients without BOS but with decreased lung function is not a surprise, given the small number of such scans in our training set. Training the DNN specifically to differentiate scans of patients with BOS from scans of a larger group of patients with confounding diseases would help to bridge this performance gap. Since our patient cohort is not large enough to reliably verify this hypothesis, we leave investigating differentiating between BOS and specific confounding diseases for a future, large-scale study.
